# Characterization of thyroid function in euthyroid and children with subclinical hypothyroidism using a multivariate homeostatic model

**DOI:** 10.1101/2023.08.26.23294665

**Authors:** Aristeidis Giannakopoulos, Alexandra Efthymiadou, Dimitra Kritikou, Dionisios Chrysis

**Author notes:** Corresponding author Aristeidis Giannakopoulos MD, PhD, Division of Endocrinology, Department of Pediatrics, Medical School, University of Patras, Rio 26504 Patras Greece.

## Abstract

**Background:** Subclinical hypothyroidism (SH) is biochemically defined by increased TSH, and normal thyroid hormones and its management is a matter of debate. Herein, we investigated thyroid function in euthyroid and children with SH using published data from population-based curves and a structure parameter inference approach (SPINA) model.

**Methods:** The study included 179 children and adolescents with SH and 311 healthy controls. The predicted and calculated secretory capacity of thyroid gland (SPINA-GT) was calculated in all euthyroid children divided into quartiles according to TSH values, and in children with SH, further subcategorized into those with mild SH (TSH: 4.5 – 10 mIU/L) and severe SH (TSH > 10 mIU/L).

**Results:** Calculated SPINA-GT values decreased significantly (P < 0.001) from the 1^st^ to the 2^nd^ quartile of normal TSH values in euthyroid children. It was also significantly decreased in mild SH compared to euthyroid children with TSH values within the upper 2 quartiles of TSH range and in severe SH compared to mild SH.

**Conclusions:** The implementation of SPINA model for thyroid function gives a wider perspective of thyroid gland’s performance within the euthyroid range of TSH, as well as in SH and add to the discussion for the long-term effects of SH and its management.

## Introduction

In current pediatric clinical practice, diagnosis, and management of typical thyroidopathies such as overt hypothyroidism or hyperthyroidism is an unambiguous task, thanks to sensitive assays for serum TSH and free thyroid hormones. However, a state of thyroid function, often encountered in pediatrics, is subclinical hypothyroidism, which is defined by normal serum concentration of total thyroxine (T4) or free T4 (FT4) and serum TSH level above the upper limit of the reference range (TSH >4.5 mIU/L).(Biondi and Cooper 2008; Biondi et al. 2019) Diagnosis of SH is usually not based on specific clinical signs or symptoms, but rather emerges as an incidental finding in a laboratory workup including thyroid function, and its management regarding therapy with thyroxine or simple monitoring, is questionable. After the neonatal period, SH is further subcategorized by serum TSH levels, in a mild (TSH 4.5 to 10 mIU/L) or a severe form (TSH > 10 mIU/L).(Biondi et al. 2019) While progression to overt hypothyroidism is possible, more commonly, stable persistence or even spontaneous normalization of the elevated TSH is observed during follow up.(Karmisholt et al. 2008)

The normal range of serum TSH levels in children and adolescents is well defined as in adults and displays a skewed distribution. (Lewandowski 2015) Serum concentrations of TSH and T4 (or FT4) are tightly regulated by the thyroid homeostatic mechanisms in a way that minor changes in circulating FT4 concentrations result in large relative changes in TSH. (Wehmann and Nisula 1984) Several studies had initially described the TSH - T4 relationship as an inverse log linear one (SPENCER et al. 1990; Meier et al. 1993; Benhadi et al. 2010), although later population-based analysis described the TSH - FT4 correlation, by 2 overlapping negative sigmoid curves with some variations attributed to age and sex differences. (Hadlow et al. 2013) Another concept of the hypothalamo-pituitary – thyroid physiology proven over the years, is the existence of the so-called set-point of thyroid function, which reflects the individual characteristics of both thyroid gland response to TSH stimulation, and feedback loop of thyroid hormones to hypothalamus and pituitary. This was based on the evidence by a study showing that individual reference ranges for serum T3 and T4 had half the width of population-based reference ranges and so the intraindividual variability of TSH and FT4 was much narrower than the interindividual one (Andersen et al. 2002). Although set points are genetically determined (Panicker et al. 2010; Walsh 2011), transposition of thyroid set point may occur with aging. (Boucai and Surks 2009; Bremner et al. 2012)

Based on the critical effect of thyroid hormones on brain development and metabolic homeostasis, the unanswered questions regarding SH in childhood are, whether the persistence of this thyroid functional state has any long-term adverse effects on CNS development,(Williams 2008; Cerbone et al. 2011; Capalbo et al. 2020) especially during infancy, or on the cardiovascular system at older ages.(Nader et al. 2010; Cerbone et al. 2014, 2016; Dahl et al. 2018) Many studies have attempted to address the above questions with contradictory results. The vagueness of the SH impact on central nervous system or cardiometabolic state and the large sample size of individuals needed to answer this question, keeps the debate on this matter active.

To investigate more in depth the nature of SH in children, we studied the thyroid function by implementing previously published population-based analytical models that describe the TSH - FT4 relationship (Hadlow et al. 2013; Fitzgerald and Bean 2016) both in euthyroid and children with SH. More specifically, we calculated the TSH values (henceforth referred as predicted TSH) of all subjects using the equations developed by population -based TSH – FT4 data. We also calculated the thyroid production capacity as approached by multiparametric analysis,(Hoermann et al. 2015; Dietrich et al. 2016) These latter models have evolved step wisely over the year, from the standard logarithmic model of thyroid homeostasis (REICHLIN and UTIGER 1967), to the more detailed multi-dimensional and non-linear relationships between TSH, FT4, T3 and their binding proteins.(Hoermann et al. 2014, 2015, 2016) Descriptively, this platform include the Michaelis–Menten kinetics for the deiodination of T4 with a time-delay model (thyroid), a negative exponential model for feedback inhibition of TSH release, and a non-linear description of plasma protein binding.(Eisenberg et al. 2008; Ben-Shachar et al. 2012) The thyroid’s secretory capacity (SPINA-GT), also referred to as thyroid output or thyroid capacity, provides an estimate for the maximum secretion rate of thyroid hormone under stimulated conditions.(Dietrich et al. 2016) The development of SPINA-GT was promoted by a long-standing reflection on non-improvement of hypothyroid symptoms of a fraction of patients that were under treatment with levothyroxine and nonetheless were rendered euthyroid biochemically.(Abdalla and Bianco 2014; Wiersinga 2014) The SPINA parameters have been validated in a number of studies in different populations comprising together more than 10,000 subjects (Dietrich et al. 2008; Hoermann et al. 2013) To the best of our knowledge, no studies have been published regarding SPINA implementation for thyroid function in children.

This detailed study of the hormonal balance of thyroid gland aim to deliver a functional insight that goes beyond the information from measuring serum TSH and FT4 levels, and to interpret the results of classical thyroid analysis under the prism of multivariate modeling.

## Methods

The study included 179 children and adolescents of Greek origin with SH with age 7.25 years (median) (range: 1 – 14.9 years) and 309 healthy controls with age 8.5 years (median) (range: 1.3 – 16,7 years). SH was defined as serum TSH > 4.5 μIU/L (min = 4.51 -and max = 15) with FT4 levels within normal range (min = 10.43 pmol/L and max = 24.5 pmol/L). The control group consisted of children and adolescents who presented to our department due to parental concern for short stature or for thyroid evaluation due to positive family history for thyroid disease. Subjects with known chronic disease, specific or non-specific clinical symptomatology or receiving any medication were excluded from the study. Informed written consent was obtained from participating individuals and their parents and was approved by the local Ethical Committee of the University Hospital of Patras, Greece. All our subjects were considered as iodine sufficient, since Greece belongs to the group of countries with adequate iodine nutrition (Zimmermann and Andersson 2021). Date of birth, family history of thyroid disorders and any acute or chronic disease were recorded. Body height and weight were measured using standard anthropometric techniques. Height, recorded to the nearest millimeter, was found as the average of two measurements using a calibrated Harpenden stadiometer. Weight was measured on a calibrated scale (Seca, model 760) and recorded to the nearest 100 gr. Pubertal stage was defined by the same investigator using Tanner’s classification. Children were classified as pre-pubertal (Tanner stage I) and pubertal (Tanner stage II to V). BMI was calculated using the formula weight/height2me (kg/m2). BMI standard deviations score (SDS) was computed for each subject using the formula: BMI-SDS = (actual BMI-mean BMI for age, race and gender)/BMI SD for age and gender.

Serum TSH, FT4, Thyroglobulin (TG-Ab) and thyroid peroxidase (TPO-Ab) antibodies were measured by electro-chemiluminescence (Elecsys 2010, Roche Diagnostics). Anti-thyroid antibody status was considered positive when TG-Ab and/or TPO-Ab was positive (>34 IU/mL).

The relationship between the natural logarithm of TSH and free T4 in subjects > 1 year old, not receiving thyroxine treatment, were described by the following equations (Hadlow et al. 2013) :

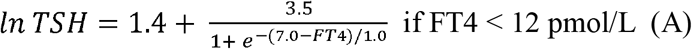

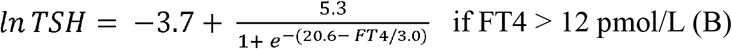

Predicted *lnTSH* was calculated using the above formulas, and results were back log-transformed to predicted TSH.

The thyroid’s secretory capacity (SPINA-GT) was defined by the formula:

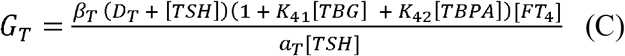

(α_T_ : Dilution factor for thyroxine = 0.1 L^-1^ β_T_ : Clearance exponent for T4 = 1.1e-6 s^-1^ D_T_ : EC_50_ for TSH = 2.75 mIU/L K_41_ : Dissociation constant of T4 at thyroxine-binding globulin = 2e10L/mol K_42_ : Dissociation constant of T4 at transthyretin = 2e8 L/mol [TBG] : standard concentration of thyroxine-binding globulin = 300nmol/L [TBPA] : Standard transthyretin concentration = 4.5 μmol/L) as a function of equilibrium concentrations of TSH, free T4, and constants or measured values for dissociation, protein binding, distribution, and elimination.(Dietrich et al. 2016) The reference range of SPINA-GT is between 1.4 and 8.7 pmol/s.(Dietrich et al. 2015)

### Statistics

Normally distributed parametric data were presented as mean ± SD and were compared with the two-sided Student’s t-test. Statistical comparison of SPINA-GT among groups defined by different TSH values was performed with analysis of variance (ANOVA). Statistical analysis of thyroid antibody positivity between groups was performed by Pearson’s chi-squared test. A p value of <0.05 was considered statistically significant in all instances. For the analysis, Stata (version 16) was used.

## Results

All 489 subjects (Male/Female ratio: 227 / 262) had an average age of 8.17 years (median) with a range of 1.1 to 16.75 years old. The SH group (n=179) was of younger age than the control group (n=309) (7.46 vs 8.6 years, p<0.05) but the BMI Z-score was not statistically different (Table 1). Analysis of thyroid function in both groups using serum TSH and FT4 measurements showed the higher TSH and lower FT4 serum levels in SH group (p<0.001) as expected. Furthermore, in SH group, TSH was negatively correlated with FT4 (r=0.26, p<0.005). Thyroid antibody positivity was statistically higher (12.3%) in the SH group (n=22/179) compared to the euthyroid group (6.4%) (n=20/313). Firstly, we used the equations (A) and (B) (see methods section) that derived from the population curves of TSH and FT4 relationship (Hadlow et al. 2013) to produce the set of predicted TSH values that correspond to the FT4 values of our subjects belonging to either euthyroid or SH group. With this approach we tried to estimate the dispersion of TSH values around the model-predicted ones for the SH group, compared to the euthyroid group. The measured TSH values against the predicted ones were plotted for both euthyroid and SH group in figure 1 (a and b respectively). We then implemented the SPINA-GT formula on all children (euthyroid and SH group), using the actual univariate TSH - FT4 paired values for each subject, and the plots of GT values according to measured TSH or FT4 are shown in Figure 2a and 2b, respectively. The correlation of SPINA-GT with TSH values of the entire range (euthyroid and SH) is graphically represented by an L-shaped scatterplot with a steep decrease within the lower range of TSH values followed by a gradual decrease of GT towards the higher TSH values (Figure 2a). Additionally, SPINA-GT values were significantly correlated to FT4 levels (r=0.523, p<0.01) as expected (figure 2b).

**Table 1.** Demographics, BMI, and biochemical indices of thyroid function in children with SH and euthyroid children.

|  | SH group | Euthyroid group |
| --- | --- | --- |
| Individuals | 179 | 311 |
| Age (years) | 7.46 ± 3.6 | 8.6 ± 3.47 |
| Sex (M/F) | 84 / 99 | 145 / 168 |
| BMI Z-score | 1.11 ± 1.46 | 1.03 ± 1.59 |
| TSH (mIU/L) | 6.7 ± 2.05* | 3.01 ± 0.95 |
| FT4 (µg/dl) | 1.22 ± 0.18** | 1.27 ± 0.17 |
| Individuals with TG-Ab (+)<br>AND/OR TPO-Ab (+) | 22 (12.3%)** | 20 (6.4%) |

\*p<0.0001
\*\*p<0.05

**Figure 1.**
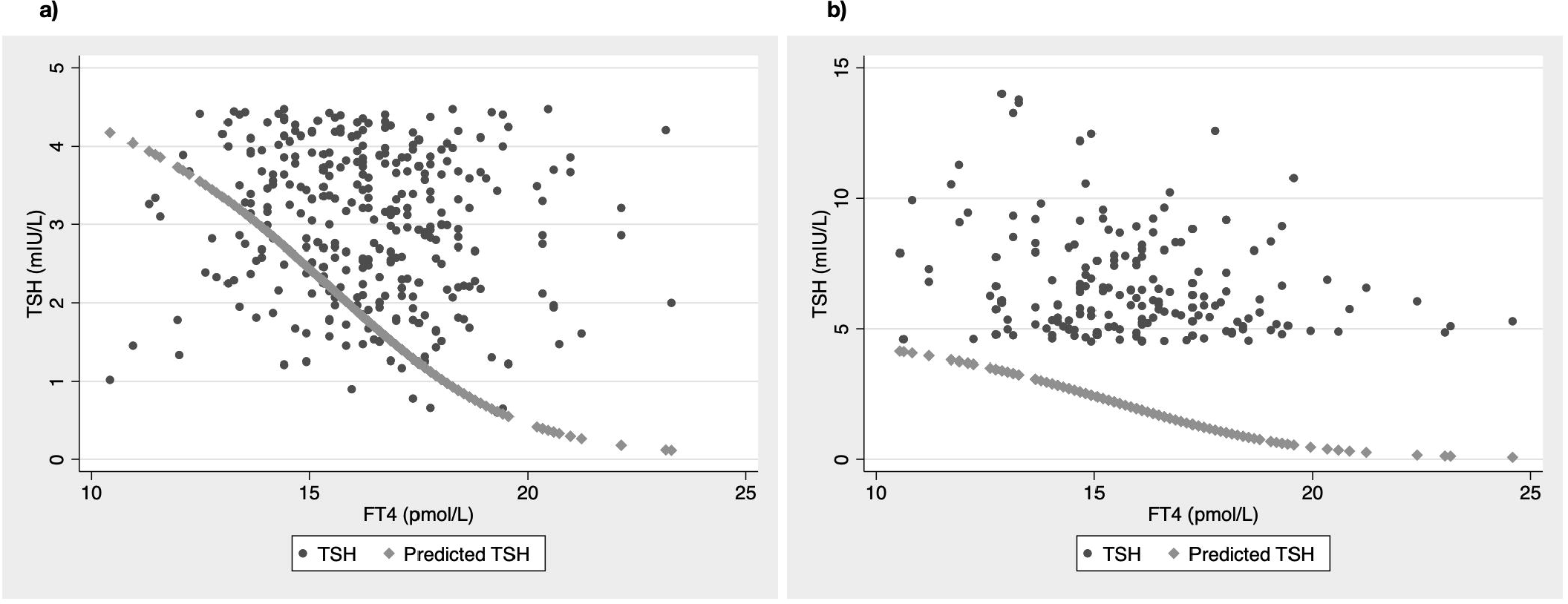
Correlation of SPINA-GT to TSH (a) and FT4 (B) in all children (SH and euthyroid)

**Figure 2.**
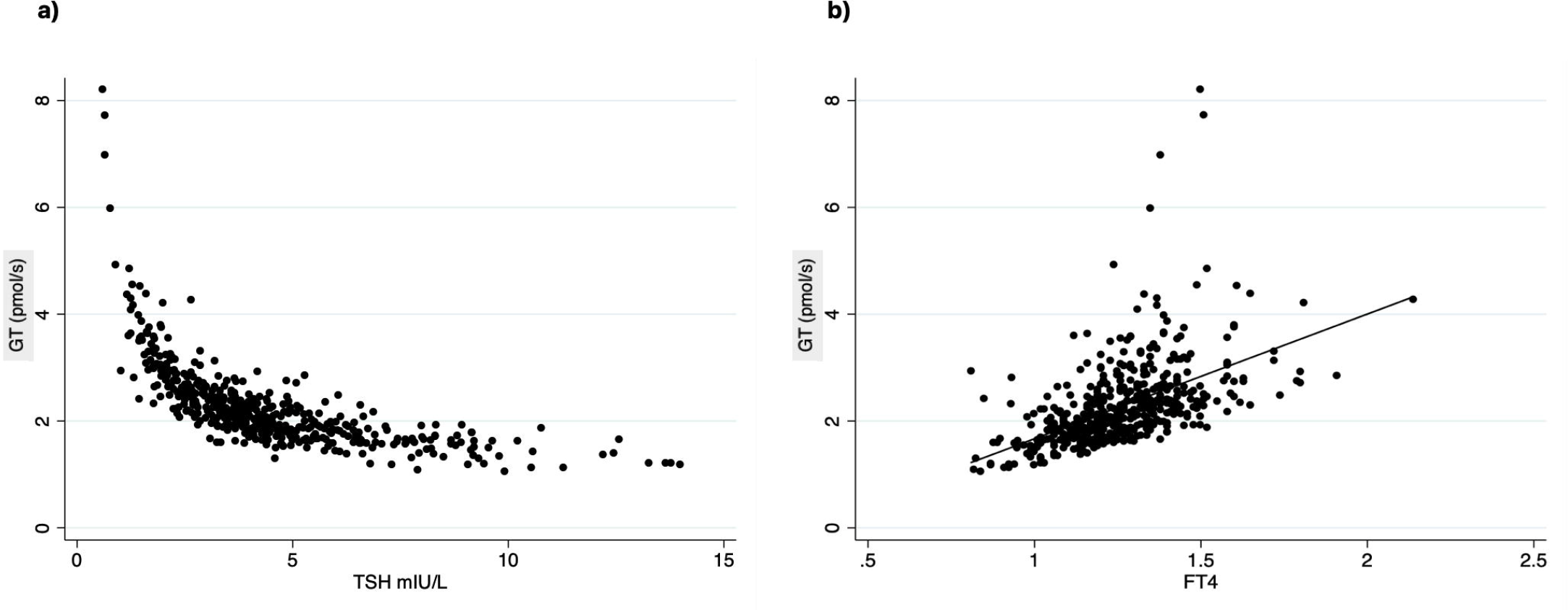
SPINA-GT values (a) in euthyroid children divided in TSH quartile groups and (b) in children with SH compared to normal TSH (upper 2 qrts) group (* P < 0.001)

Subsequently, we analyzed the SPINA-GT levels in euthyroid children divided in 4 groups of TSH values according to TSH quartiles and in children with mild and severe SH. We found that within the euthyroid range of TSH values, SPINA-GT was significantly lower in the 2^nd^ quartile compared to the 1^st^ quartile (p<0.001) while the lower values of SPINA-GT seen in the 3^rd^ and 4^th^ TSH quartiles were not statistically different compared to the 2^nd^ quartile (p values (figure 3a). We also compared the above calculated SPINA-GT levels to the predicted SPINA-GT levels that derived from the predicted TSH values (calculated from the equations A and B that were described in methods) instead of the actual ones. The intraindividual difference was statistically significant in all quartiles (p <0.01 for the 1^st^ quartile and p < 0.001 for the 2^nd,^ 3^rd and 4th^ quartile. (Figure 3a) Next, we studied the calculated and predicted SPINA-GT profiles in the upper two TSH quartiles of the euthyroid children, in children with mild SH, and in children with severe SH. SPINA-GT levels were statistically lower in children with SH than in the euthyroid group with TSH levels in the upper 2 quartiles (p <0.001). Respectively, the group with severe SH had statistically lower levels than children with mild SH (p<0.001) (figure 3b). In subjects of the euthyroid group with TSH values in the upper 2 quartiles and in subjects of the SH group with mild or severe SH the predicted GT values were all significantly higher compared to the calculated GT values (p<0.001 for all groups). Regarding thyroid antibody positivity, individuals within the upper 2 quartiles of the euthyroid TSH range and SH show increased positivity in thyroid antibodies compared to the lower 2 quartiles of the euthyroid TSH range although without any statistical significance. (Table 2)

**Table 2.** TSH values (min and max) in euthyroid (by quartiles) and children with SH and their corresponding SPINA-GT value (median and mean) along with frequency of thyroid antibody positivity.

| Euthyroid<br>(Quartile) | TSH<br>(min) | TSH<br>(max) | SPINA-GT<br>(median) | SPINA-GT<br>(mean) | Individuals with positive<br>thyroid antibodies |
| --- | --- | --- | --- | --- | --- |
| 1<sup>st</sup> | 0.6 | 2.25 | 3.21 | 3.49 | 3.7% (3/80) |
| 2<sup>nd</sup> | 2.26 | 3.14 | 2.53 | 2.52 | 3.9% (3/77) |
| 3<sup>rd</sup> | 3.15 | 3.82 | 2.20 | 2.22 | 10% (8/78) |
| 4<sup>th</sup> | 3.85 | 4.47 | 1.70 | 2.03 | 7.7% (6/78) |
| Mild SH | 4.5 | 9.93 | 1.77 | 1.74 | 12% (20/171) |
| Severe SH | 10.23 | 14 | 1.29 | 1.36 | 17% (2/12) |

**Figure 3.**
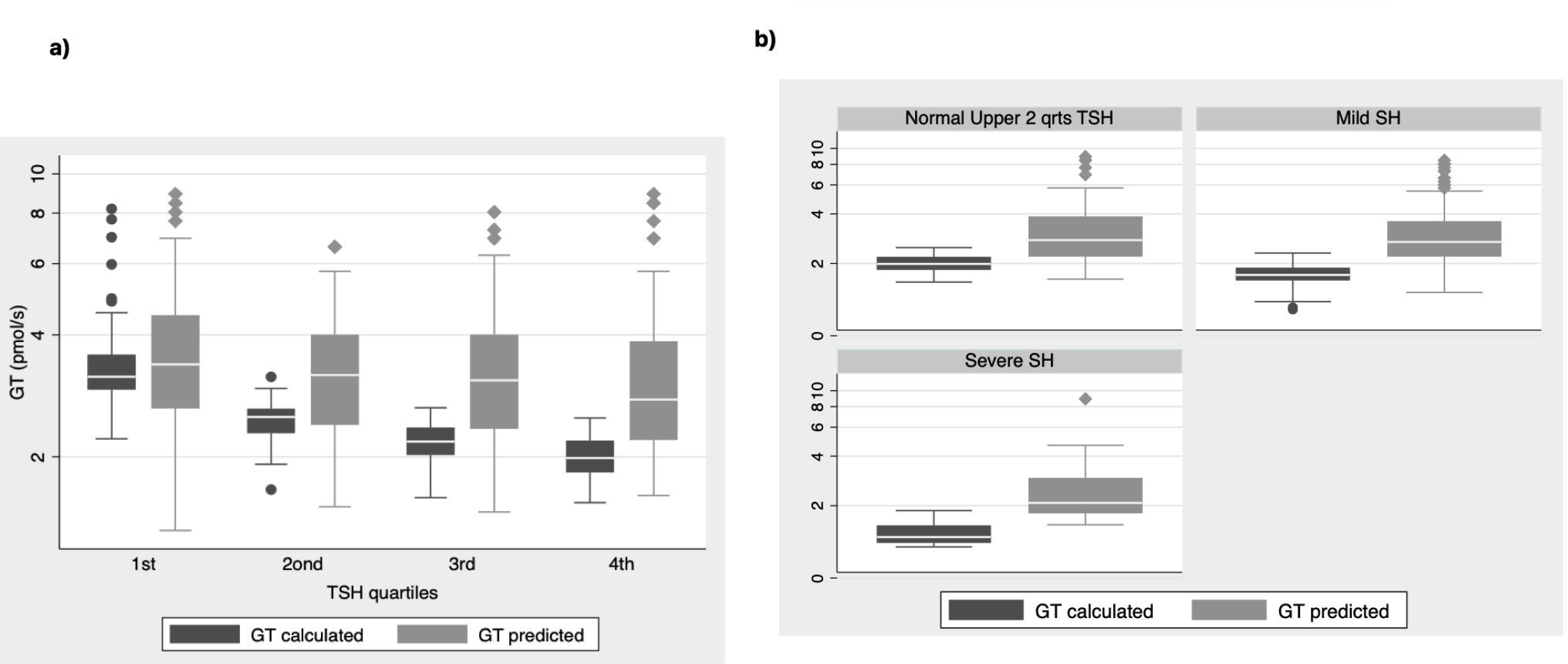
a) SPINA-GT calculated vs predicted values in a) euthyroid children and b) in children with SH compared to the upper 2 TSH quartiles of euthyroid range.

## Discussion

Subclinical hypothyroidism in children, a biochemically defined condition with no clinical symptomatology, has always been a puzzling entity regarding the biological significance and the need for treatment. The debate on the necessity of therapy in SH stems mainly from its probable long-term implications on neurodevelopment, given the effect of thyroid hormones on pre and postnatal brain development(Horn and Heuer 2010) and the documented mental impairment caused by overt hypothyroidism before 3 years of age.(Collaborative 1985; Counts and Varma 2009) The worse neurocognitive outcome found in school-age children whose TSH values from neonatal screening were between the 99.5th and 99.9th percentiles by Lain et al.(Lain et al. 2016), in a population study with significant statistical power, certainly argues in favor of treating SH in very young ages. However, other but smaller studies do not support this finding.(Cerbone et al. 2011; Trumpff et al. 2016) In cardio-metabolic health, higher lipid levels and more specifically non-HDL C, triglycerides and total cholesterol have been found to be statistically higher in SH by many studies.(Nader et al. 2010; Cerbone et al. 2014; Dahl et al. 2018; Jin 2018) On the contrary, blood pressure (Ittermann et al. 2012; Lee et al. 2019) and glucose metabolism (Cerbone et al. 2014; Jin 2018) have not shown any consistent correlation with the increase of TSH in the pediatric population.

Our current estimation of thyroid function is based on measuring TSH and FT4 in serum. Earlier studies on thyroid homeostasis, described by measuring TSH – FT4 or T3 pair values concluded that the intraindividual variations of serum TSH, FT4 and FT3 are very narrow. (Harrop et al. 1984; Andersen et al. 2002) Additionally, the increased variability of the working point of HPT axis in healthy individuals, (Andersen et al. 2002) limits the usefulness of population-based reference ranges in the identification of a disease state.(Benhadi et al. 2010) Consequently, measured values of TSH and FT4 within the normal range, does not necessarily indicate a normal thyroid function for the specific individual. This is also reflected in the differential increase of FT4 to stimulation by TSH, or TSH suppression secondary to T4 increase, where differences in thyroid physiological processes or in thyroid gland size may have a role. (Fitzgerald and Bean 2016). In agreement with the data, we saw a large dispersion of TSH values around the fit line of predicted TSH based on the population-based model in euthyroid children showing the significant variation of HPT set points in euthyroidism. However, in children with SH, all TSH values were placed above the fit line of the predicted TSH, as expected (Figure 1).

Beyond the above-mentioned unresolved issues on the biological consequences and treatment in children with SH, more questions also arise about the validity of thyroid evaluation using the current univariate method of measuring TSH and thyroid hormones in serum, used in everyday practice. This is supported by the discordance of clinical symptoms to biochemical indices of thyroid physiology (Abdalla and Bianco 2014; Wiersinga 2014) as reported by some patients expressing fatigue and not-wellbeing under treatment with thyroxine and biochemically restored euthyroidism.(Hoermann et al. 2015, 2016) Consequently, existing multivariate models of thyroid homeostasis that have been developed and incorporate much of the complexity of the feedback systems in thyroid hormonal production, have been applied in the study of thyroid function. In the case of SH, to further investigate whether the modestly increased TSH stands for a chronic compensated state of thyroid function, or it is a simple functional and possible transient transposition of the thyroid gland’s homeostatic set point, we used the SPINA-GT model. The *in vivo* evaluation of this model in adults has proven that it can clearly differentiate between the primary thyroid disorders and euthyroidism, and it is not affected by the TSH values. (Hoermann et al. 2016) This is important for the evaluation of SH since it is a primary thyroid dysfunction detected only by the increased TSH levels. SPINA GT model is implemented for the first time in the pediatric population and the constants TBG and TBPA, used in the model’s equation have the same values in children. (Neto and Rubin 2001; Espe et al. 2007). From our results, it can be easily observed that thyroidal secretion rate (SPINA-GT) follows an L-shaped curve which presents with a steep depression within the euthyroid range of TSH values and then displays a lesser degree of decrease at TSH levels over 4.5 mIU/L. Regarding only the euthyroid subjects, we can see that thyroid secretion rate is statistically different only between the 1^st^ and 2^nd^ quartile of TSH values of euthyroid range. Comparison among the 2^nd^, 3^rd^, and 4^th^ quartiles of TSH do not display any statistical difference (Figure 2a). By comparing the calculated SPINA GT values to the predicted SPINA-GT values, we see the same differences with significant differences in thyroid production between the 1^st^ and 2^nd^ TSH quartile (Figure 2a) and between the upper 2 quartiles of TSH in euthyroid group and children with SH (Figure 2b).

The above observations first point to the concept of diverse thyroid set points within the euthyroid range supporting a different secretion potential across euthyroidism. The lower secretion capacity of thyroid hormones within the euthyroid range of normal healthy children is revealed only by the SPINA-GT model and not appreciated by the classical univariate analysis. The side-by-side analysis of SPINA-GT calculated values and predicted values show that predicted SPINA-GT values (based on the population-based predicted TSH values) do not have any statistical differences within the TSH-euthyroid range (Figure 3a). Within the SH TSH range (TSH > 4.5 mIU/L), SPINA-GT values are significantly lower compared to the upper 2 quartiles of normal TSH. Additionally, severe SH (TSH> 10 mIU/L) has significantly lower SPINA-GT levels from mild SH. In parallel, within the SH range SPINA GT predicted values are significantly increased compared to SPINA-GT calculated values. (Figure 3b) These observations are compatible to a transposition of the set point line (Figure 4) in thyroid homeostasis in SH which we do not know its significance on the clinical level. A more thorough evaluation of thyroid function in this case, should also include the link of secretion dynamics to the sum activity of peripheral deiodinases which assesses the thyroid action on tissues. A parallel increase of type 2 deiodinase activity at the tissue level could compensate for the lower central thyroid production of FT4. The latter effect is measured by the SPINA-GD model by T3 as a variable. (Dietrich et al. 2016) This is a limitation of our study since we did not have T3 measurements in our subjects. However, FT4, as a factor that takes part in the homeostasis of both secretion and deiodination, reflects the connection between thyroid production and feedback. In addition, it has been argued that the pituitary gland is more responsive to T3 generated in the pituitary from the circulating T4 by the action of type 2 deiodinase, than to circulating T3.(Goede et al. 2014b) In support to the above is the more consistent correlation of TSH to FT4 rather than T3.(Goede et al. 2014b, 2014a) The continuum of lowered thyroid hormone production in SH may raise questions regarding any possible consequences on brain development in infants.

**Figure 4.**
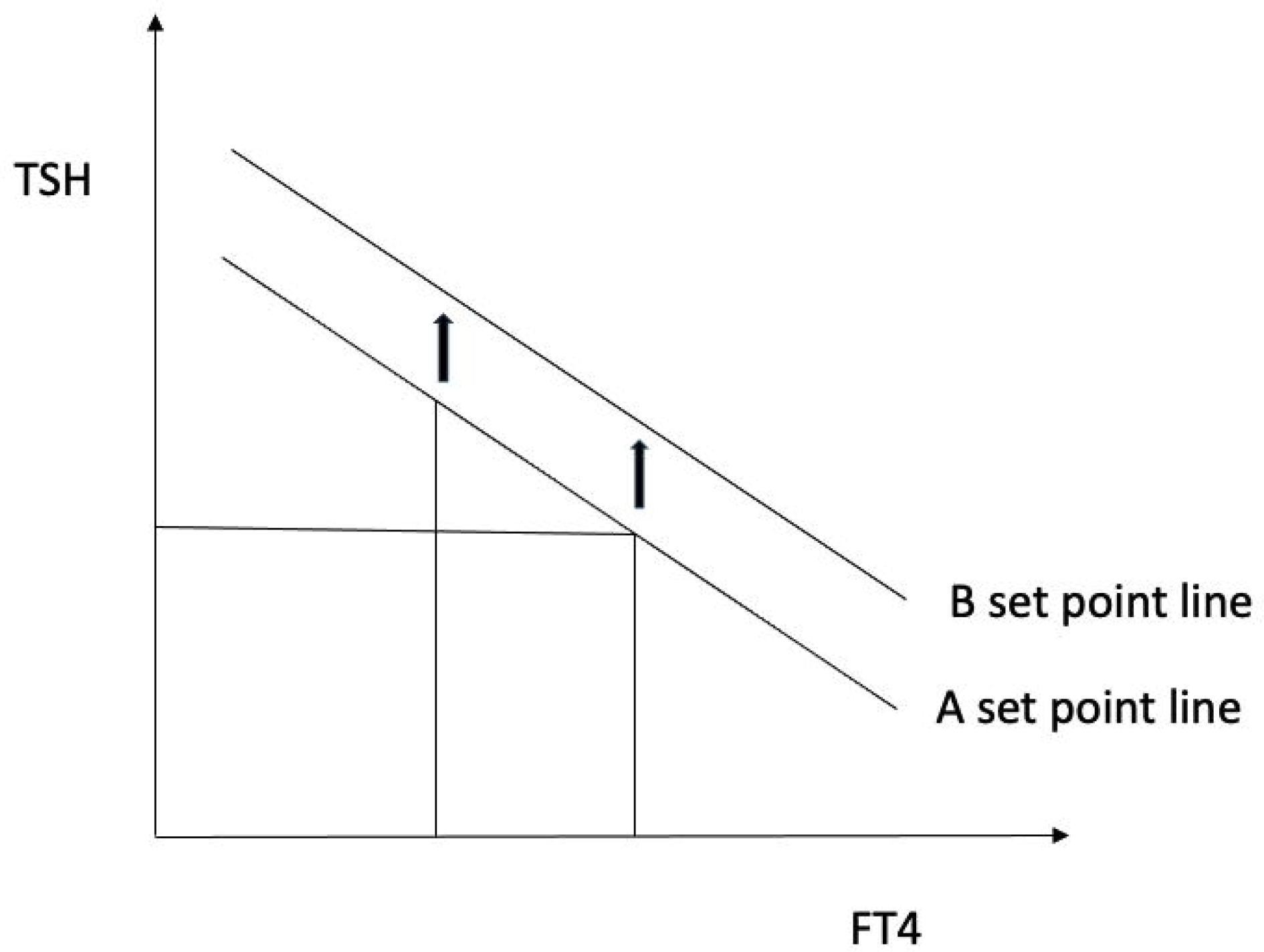
Schematic diagram depicting the transposition of set point line (from A to B) in SH across a small range of FT4 values.

While the alterations in thyroid production in SH may be interpretable, the interesting point in the drop of SPINA GT value in the 1^st^ to 2^nd^ quartile of TSH in the euthyroid range. While we cannot attribute any physiological meaning in this change, it should be interpreted in association with the SPINA-GD results, in which increased levels of SPINA GD could compensate for the lower SPINA-GT levels and result on the same fingerprint on thyroid hormone action. Otherwise, a lower thyroid hormone output within the euthyroid range could append the conversation of the current TSH upper reference limits in children and support a further skewed to the left distribution of TSH values. This argument has been proposed by a study that supports that TSH reference distribution may be skewed by an occult thyroid dysfunction based on increased thyroid antibodies found in the adolescent group (12-19yr) when they presented with TSH values > 2,5 mIU/L.(Spencer et al. 2007) In our study, we found increased thyroid antibodies in the 3^rd^ and 4^th^ quartile of euthyroid TSH values which may imply of the existence of an occult thyroid disease in a small percentage of children (Table 2) but had no statistical significance compared to the first 2 quartiles. Of note is, that the upper 2 quartiles of TSH levels of euthyroidism and SH showed similar percentages of thyroid antibody positivity.

## Conclusions

The implementation of multivariate models such as SPINA-GT for thyroid function gives us a wider perspective of thyroid’s functional status within the euthyroid range of TSH, as well as in the state of SH and supply more information in the interpretation of the classical univariate thyroid analysis. The above thyroid homeostatic changes need to be confirmed by further studies prior to discussion of their meaning for its long-term effects on health and their potential for medical decision making.

## Data availability statement

The datasets analyzed during the current study are available from the corresponding author on reasonable request.

## Funding

No funding was used for this study.

## Authors’ contribution

AG conceived, analyzed the data, and drafted the manuscript, AE and DK collected the samples, DC conceived and reviewed the manuscript.

## Competing interests

The authors have nothing to disclose.

## Notes

### Competing Interest Statement

The authors have declared no competing interest.

### Funding Statement

This study did not receive any funding

### Author Declarations

Ethics committee of University Hospital of Patras, Greece approved the protocol.

